# Enhancing Research Data Infrastructure to Address the Opioid Epidemic: The Opioid Overdose Network (02-Net)

**DOI:** 10.1101/2021.07.15.21260134

**Authors:** Leslie A Lenert, Vivienne Zhu, Lindsey Jennings, Jenna L McCauley, Jihad S Obeid, Ralph Ward, Saeed Hassanpour, Lisa A Marsch, Michael Hogarth, Perry Shipman, Daniel R Harris, Jeffery C Talbert

## Abstract

**Objective:** Opioid Overdose Network is an effort to generalize and adapt an existing research data network, the Accrual to Clinical Trials (ACT) Network, to support design of trials for survivors of opioid overdoses presenting to emergency departments (ED). Four institutions (Medical University of South Carolina (MUSC), Dartmouth Medical School (DMS), University of Kentucky (UK), and University of California San Diego (UCSD)) worked to adapt ACT network. This paper reports their progress.

**Materials and Methods:** The approach taken to enhancing ACT network focused on four activities: cloning and extending the ACT infrastructure, developing an e-phenotype and corresponding registry, developing portable natural language processing (NLP) tools to enhance data capture, and developing automated documentation templates to enhance extended data capture.

**Results:** All four institutions were able to replicate their i2b2 and Shared Health Research Information Network (SHRINE) infrastructure. A five category e-phenotype model based on ICD-10 coding was developed from prior published work. Ongoing work is refining this via machine learning and artificial intelligence methods. Portable NLP tools, focused at the sentence level, were also developed to identify uncoded opioid overdose related concepts in provider notes.

Optimal performance was seen in NLP tools that combined rule-based with deep learning methods (F score, 0.94). A template for ED overdose documentation was developed to improve primary data capture. Interactive prompts to physicians inside ED progress notes were effective in promoting use of the template. The template had good system usability and net promoter scores (0.72 and 0.75, respectively, n=13). Work to design ED trials based on the network’s data is underway.

**Discussion and Conclusions:** Overall, initial results suggest that tailoring of existing multipurpose research networks to specific tasks is feasible; however, substantial efforts are required for coordination of the subnetwork and development of new tools for extension of available data.

## INTRODUCTION

This paper describes work to adapt an existing nationwide clinical data network to better assess and conduct trials to combat the opioid overdose (OOD) epidemic. Opioid misuse and dependence continue to be a significant and growing cause of preventable morbidity and mortality in the United States [1]. Over the last 10 years, the number of individuals presenting to emergency departments (EDs) as a result of intentional or unintentional opioid-related overdose(OD) has dramatically increased [2][3]. Many of these individuals die or have a repeat OD presentation within a year [4][5] and public health costs associated with opioid-related OD are high [2][6][7]. The defining features of the opioid epidemic have changed over time. While prescription opioid misuse was initially a major contributing problem, increasingly the use of high potency non-prescription drugs, such as fentanyl, heroin, and illicit fentanyl have been the cause of opioid-related ED presentations and mortality [3][8]. Recent data suggest that a growing proportion of opioid-dependent individuals are initiating use with heroin (or illicit synthetic opioids), rather than prescribed opioids [9][10], contributing to public health experts’ beliefs that opioid-related OD may continue to increase over the next 1-5 years [9][10].

Patients presenting with OOD to EDs represent an extraordinarily high-risk group for mortality (up to 24-fold risk ratio) [11], and thus are a high-value target for interventions to combat the opioid epidemic. The ability to characterize these individuals could inform trials of multi-level, multi-system OD prevention efforts, as well as translational research efforts to connect opioid-dependent individuals with effective treatments.

Recognizing that accuracy and detail in preliminary data is essential when designing effective clinical and translational studies (research-driven, interactive access to data is preferable through tools such as i2b2 [12]), we have undertaken foundational work to extend the Accrual to Clinical Trials (ACT [13]) network to create an inter-institutional research database and network focused on accelerating clinical and translational research (CTR) for opioid-use disorder in the context of OOD. ACT is a network supported by The National Center for Advancing Translational Sciences (NCATS) that includes approximately 60 Clinical and Translational Science Award (CTSA)-funded sites. It has a standardized clinical data model. Through use of Shared Health Research Information Network (SHRINE) software [14], it has the unique capability of providing counts of patients meeting specific criteria, in real time, across the network. Our work predates efforts also discussed below on adapting CTSA data networks to COVID-19 research as part of NCATS National COVID Cohort Collaborator (N3C [15]) but also illustrates how this new investment might be broadened to address other national priorities for research.

To develop a test bed for adapting the ACT network to OUD and OOD scenarios, we worked with three network partners: the University of California San Diego (UCSD), the University of Kentucky (UK) and Dartmouth CTSA sites. To create this network, we developed an (1) e-phenotype for case identification in the ED based on (electronic health record) EHR data, combined this (2) with natural language processing (NLP) algorithms to obtain additional data from clinical notes, and (3) EHR tools for direct entry of data during clinical care in the ED. These tools, designed in the context of cross institution collaboration, allow for a more thorough characterization of individuals presenting to EDs with OOD including: demographics, comorbidities, OOD agent and concepts that are often incompletely represented in ED notes such as source of the opioid, intentionality of OOD, and ED treatment and discharge disposition. Our goal is to create tools and training materials that extend modifications from the initial set of institutions to the entire ACT network (60 or so CTSA institutions), so that the entire network can be used for trial planning and surveillance.

## METHODS

The goal of this work was to demonstrate the feasibility of adapting existing data resources to new and urgent tasks through enhancements of data capture. Overall design of the network is shown in Figure 1. Critical components include an EHR system, a data warehouse that is fed by the EHR, NLP processing tools, a dedicated instance of i2b2 linked to the data warehouse for OOD queries, and a SHRINE node for receipt and processing of remote data queries. Data are routed to the OOD instance from the generic research i2b2 based on an e-phenotype for eligibility. Data are enriched in two ways: 1) NLP processing to identify concepts stated in text but not encoded during case abstraction; and 2) through use of a documentation templates for the EHR that enhances the amount of coded data in the EHR and reminds providers to discuss critical concepts, such as the intentionality of the overdose.

**Figure 1.**
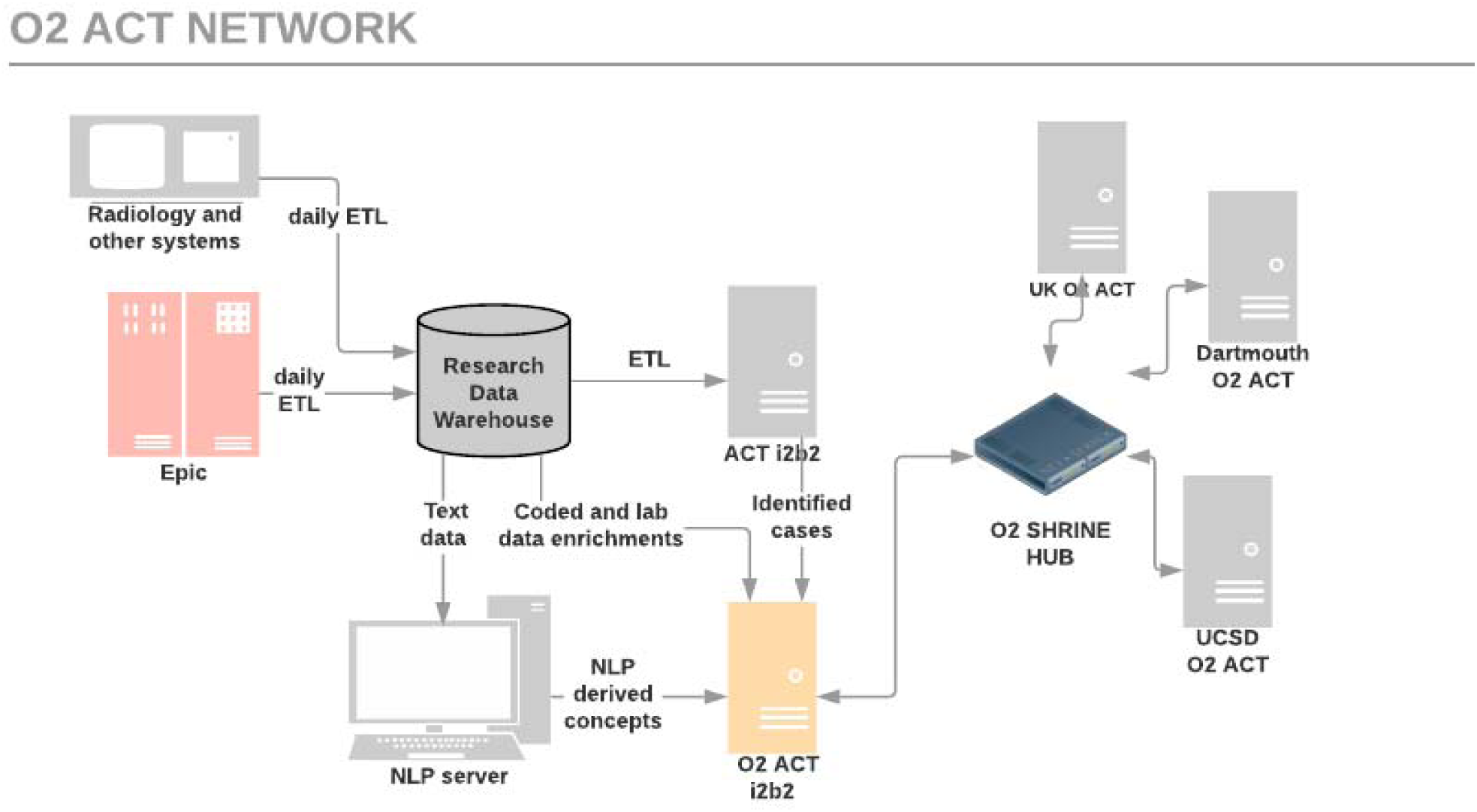
Overall architecture of O2-NET.

### Network Infrastructure

The O2-Network was designed around a parallel implementation of i2b2 and SHRINE to existing data assets. We chose this approach to allow rapid testing and adaptation of the network for critical issues. The network was anticipated to be in a development phase for several years, with the eventual goal of merging innovations into production i2b2 and SHRINE environments for NCATS through appropriate governance processes. Sites were required to clone and install a second ACT i2b2 instance and SHRINE node for this purpose.

### E-phenotype Definition

The ICD-10 and laboratory data-based e-phenotyping team was led by an OUD researcher. The primary objective of this team is to develop precise OOD cohort entry defictions. This team’s approach was focused towards harmonization of prior literature on definitions of OOD published in the literature. This team conducted a scoping review [16] of the literature on ICD-driven phenotypes for identification of OOD using PubMed search with snowball sampling of the literature cited by discovered papers. Two main categories of studies were included in the review: (1) those that developed or evaluated an e-phenotype for opioid-related OD; and (2) those that employed a case definition for opioid-related OD as a means of identifying EHR records for cohort inclusion. The literature review was updated through October 2019. Preliminary studies suggested that a tiered framework for representation of the probability of a true case could be important. High probability cases would probably have an OOD specific ICD-10 code at the time of case abstraction. Medium and low probability cases would be OOD cases that were not correctly coded or had some other medical problem that superseded the OD in retrospective coding and/or physician billing codes. Based on the literature and expert knowledge, we created a framework for categorizing cases according to their probability based on ICD-10 codes, medication codes, and laboratory codes with five different sub-types.

To validate the framework, assess variability in coding, and support refinement of the e-phenotype using machine learning (ML) approaches, we are creating a gold standard data set. ED visit clinical notes classified into “at risk” or OD events, based on an early ICD-10 based case definition, were exported into a REDCap application. Remote and in person reviewers used this application to annotate notes with a document-level classification of the probability of the case representing an OOD event. Our aim is to review around 2250 cases, based on training set size desired for ML and sample size calculations for precision of estimates. A team of twelve reviewers (clinicians, clinical psychologists, and medical students) was assembled for this purpose and their work is ongoing.

### Application of NLP, Machine Learning and Artificial Intelligence Methods

The primary goal of the NLP team is to enrich data in identified cases with critical concepts for OUD and OOD identified in the notes. The NLP team was led by an experienced informatician and its objectives and work reflected that perspective. *Their approach was focused on abstraction at the sentence level of medical facts* about the case (as opposed to the other team’s work on classification of cases for cohort generation.) We believed many key features of extant cases of OOD would be “locked” in notes of providers caring for patients in the ED. Examples of data that might only be recorded in provider notes include level of consciousness on admission, use of opioids in the field by paramedics, type of opioid suspected in the OD, and intentionality of the OD. We envisioned two types of information that might be abstracted from a note for OD case identification: 1) mention of OOD; and 2) documented response to OOD treatment (“Narcan” or “naloxone”). Other attributes about the case may or may not be stated in a note, but be critical to understanding context, and potentially be inferred from the note using ML and/or artificial intelligence (AI) methods. A critical detail might be inference of the intentionality of the OD. Was this a patient with chronic pain who accidentally used too much medication, an individual with OUD who unanticipatedly took a more concentrated aliquot of drug in a recreational context, or was this a suicide attempt? While such inferences might be highly precise, they may provide valuable information for trial planning and clinical intervention development. Additionally, we examined the value of AI methods such as ML and deep learning (DL) approaches in identification of OD cases.

Our initial work evaluated several different approaches to concept identification that combined ML and NLP methods using the CLAMP platform [17]. The study cohort included patients who were age 12 or older with an MUSC ED visit between 2013 and 2018 without the occurrence of a surgical procedure or cancer diagnosis during this period. Selected clinical note types included ED notes, ED follow-up notes, progress notes, discharge summaries, and consultant notes covering 99% of clinical notes for this cohort. Four NLP approaches were developed: 1) Named Entity Recognition (NER) + Rules2) A Support Vector Machine (SVM) classifier based on java libsvm library, with unigrams, bigrams, and tri-grams as a feature; 3) A Neural Network approach based on pre-trained context embeddings: Bidirectional Encoder Representations from Transformers (BERT) was used for the classification task; and 4) BERT+ Rules. Modifiers such as OD for opioid, response to Narcan, subject, condition, hypothesis, or intentionality were detected for two primary entities (opioid and Narcan). Inference rules were applied to further eliminate false positives, including situations such as the subject is not the OD patient, an OD event did not happen (e.g., condition, hypothesis), or a negation (e.g., “no,” “deny’). NLP performance was evaluated using the gold standard (manual review of potentially relevant sentences identified by Ruta). Sentences were reviewed independently by two experts (coauthors LJ and JM) with discussion to review conflicts in classification. Precision, recall, and F-score were calculated.

### EHR Modifications

While we were optimistic about the potential role of NLP and other tools in enhancing available data from patients previously treated for OOD, we believed it was preferable for incident cases to improve provider documentation of the OOD during clinical care. To achieve this goal, we worked with physicians and OUD experts to develop an enhanced template for documentation of provider notes in the ED so the necessary data for trial planning would be captured. The team was led by an ED clinician with a research focus on OUD. The goal was to produce a clinically usable tool to enhance data capture for the OOD clinical context.

To make the additional documentation for the research database more palatable for providers, the OOD template or opioid smart tool (OST) was designed to automatically produce a text note for the clinician by selecting from alternatives or filling in specific blanks. We explored various options for data capture enhancement, including having specific note templates that providers could select from a menu and shortcut tools that could be put into documentation with keywords (“.phrases”, “dot phrase”, or “smart phrase” in the EHR (Epic) developers parlance).

After a discussion with the EHR and ED teams, we implemented a dot phrase (to insert the opioid documentation tool), as it did not interfere with previously created templates for opioid documentation and was easily accessible. User-friendly features to pull in previously documented data points, such as vital signs and the Glasgow Coma Score (GCS), were also utilized. An example of the OOD template is shown in Figure 2.

**Figure 2.**
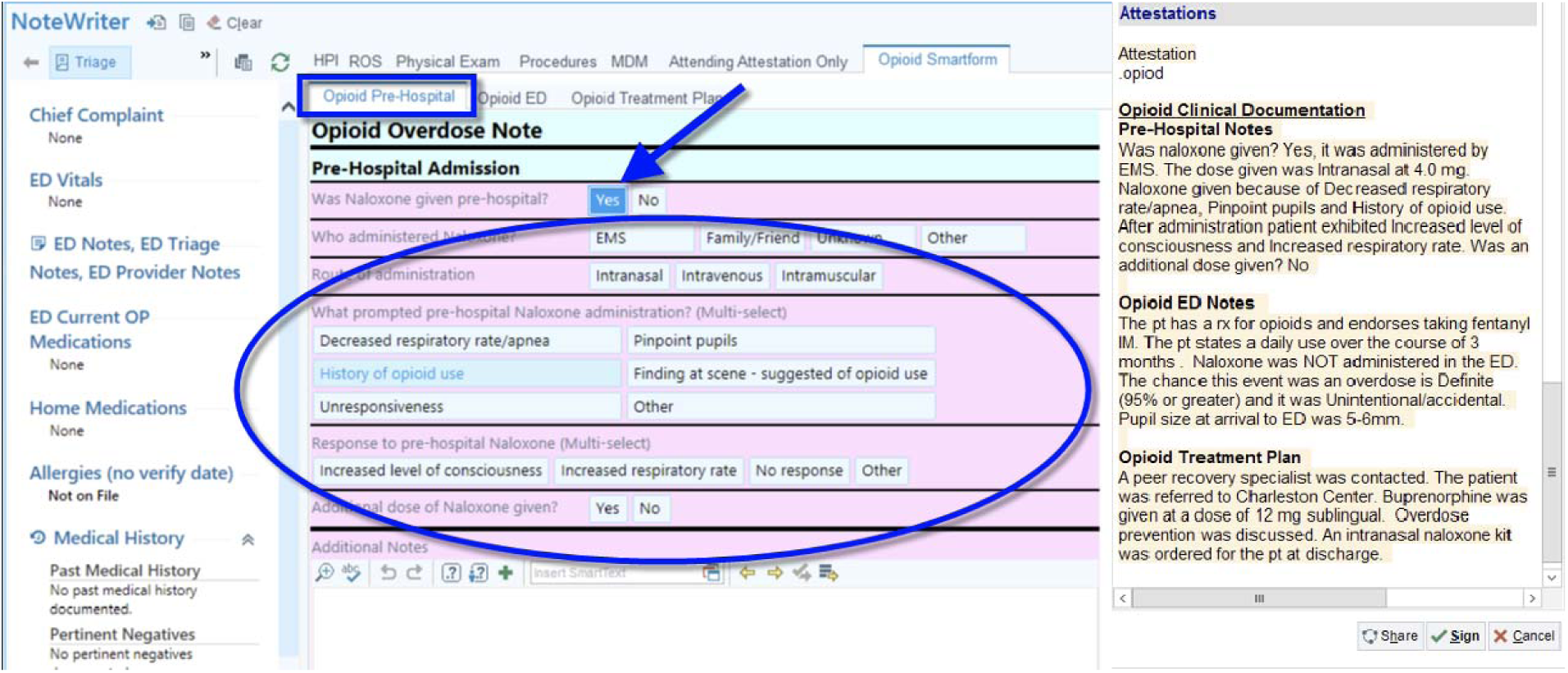
Template for documenting an opioid overdose episode and resulting provider note. Screen image used with permission from the electronic health record vendor (Epic).

We recognized that providers might not *remember* to use an unobtrusive documentation tool. Therefore, a novel approach was developed where, upon admission, a chief complaint with the words “Drug Overdose”, “naloxone”, or “Narcan” triggered the insertion of a “reminder” phrase into the history section of the ED provider’s note. Figure 3 Shows this reminder in context. Providers had to delete this text to close their progress note.

**Figure 3.**
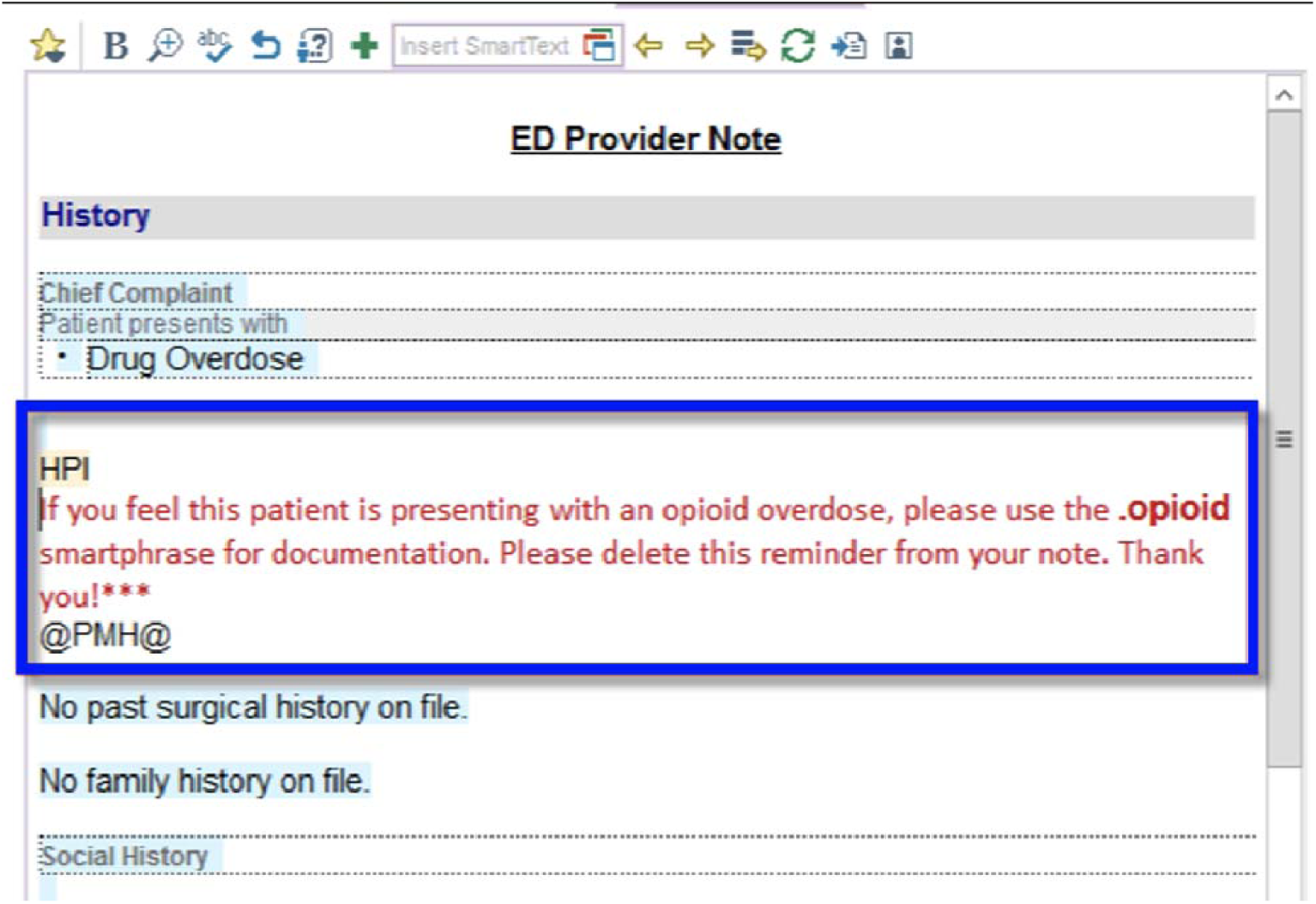
Reminder notice inserted into emergency department notes as it appears in the electronic health record (Epic). Screen image with permission from the Epic vendor.

We trained ED providers and residents in the use of the OST, initially without the reminder, with in-service meetings and creating training materials for use of the documentation tool. After a period of observation, we introduced the reminder system to examine its additional effects on utilization of the template. After achieving adequate uptake of the documentation tool, we assessed usability and provider satisfaction through the System Usability Scale (SUS) and Net Promoter Survey (NPS), respectively utilizing an anonymous survey sent to providers who had leveraged the template.

## RESULTS

Table 1 shows progress of the O2 Network at the end of year two. As shown in the table, there was substantial advancement not only with development of OOD tools but significant work on implementation. All remote sites completed initial queries of coded e-phenotypes, deployed i2b2 and SHRINE instances, initiated deployment of NLP pipelines, and started to seek governance approval of ED EHR template tools after identification of clinical champions and approval of installation by those champions.

**Table 1.** Progress on creation of O2-Net Across the consortium (counts rounded to the nearest tenth for confidentiality) based on ICD codes. Table highlights differences in case make-up.

|  | Near certain cases count (e-phenotypes 1&2) | Probable case count (e-phenotypes 3&4) | Possible case count (e-phenotype 5) | i2b2 for O2 Net | SHRINE | NLP pipeline | ED Template |
| --- | --- | --- | --- | --- | --- | --- | --- |
| MUSC | 12,230 | 3,270 | 4,080 | Operational | Operational | Operational | Deployed |
| Dartmouth | 1,580 | 280 | 260 | Operational | Operational | Deploying | Seeking IT governance approval |
| UK | 15,250 | 18,140 | 14,060 | Operational | Operational | Deploying | Seeking IT governance approval |
| UCSD | 8,800 | 6,740 | 18,310 | Operational | Operational | Deploying | Seeking IT governance approval |

### Network Deployment

Recreating dedicated i2b2 instances with ACT ontology and a secondary SHRINE network proved to be a feasible approach but required slightly more effort than first anticipated. Instances with test data were deployed at the four sites, however database engine differences (Microsoft SQL Server and Oracle DBMS) generated some configuration considerations. Firewall rules were heavily dependent on local IT security and administration, and a major SHRINE version upgrade was completed, but not anticipated. Inception to final successful site connection was achieved after about 9 months of efforts.

### E-phenotype Development

The initial step of the e-phenotype development process entailed a thorough, but non-systematic, review of the published literature, indexed with PubMed, for prior work on ICD-driven phenotypes for identification of opioid-related OD in ED settings. Two main types of studies were included in the review: (1) those that aimed to develop and/or evaluate an e-phenotype for opioid-related OD; and (2) those that employed a case definition for opioid-related OD to identify EHR records for inclusion in their study. This literature review was updated through October 2019. We subsequently compiled an exhaustive list of all ICD-9 and ICD-10 codes used in previous e-phenotypes. Of note, ICD-9 codes constituted the majority of elements from previous studies. Given the full transition of sites to ICD-10 use, all ICD-9 elements were translated into their corresponding ICD-10 codes. Given that ICD-9 to ICD-10 translation expanded the total code count substantially, the resulting ICD-10 code list was reviewed by multiple team members with clinical and topical expertise to remove ICD-10 codes with no potential relevance to opioid-related OD. Reviewed studies, as well as key information abstracted including ICD codes, are presented in Supplemental Table 1.

Based on guidance from previous research, MUSC assembled a team of multidisciplinary clinical experts representing the domains of addiction science, emergency medicine, bioinformatics, and health services research. Consistent with the commonalities of prior work in this area, *a priori* case definitions developed by the lead team included two key components: (1) some indication of opioid use/involvement; and (2) some indication of respiratory suppression and/or loss of consciousness. A *priori* definitions were reviewed and approved by clinical experts at each of the three partnering national CTSA sites (UK, UCSD, and Dartmouth).

This work resulted in five categories of e-phenotypes, each reflecting a varying degree of certainty that the defined criteria would yield case inclusion with varying degrees of targeted positive predictive value (PPV). These five categories were: (1) definite opioid-related OD via administrative coding; (2) definite opioid-related OD via coded naloxone administration and coded positive response; (3) probable opioid-related OD via documented naloxone administration in the context of diagnosed or suspected OUD; (4) probable opioid-related OD via other indicators in the context of diagnosed or suspected OUD;, and (5) possible opioid-related OD. The proposed a *priori* case definitions for each category and associated ICD-10 codes resulting from this process are presented in Supplemental Table 2.

Case counts were requested from each site for the number of ED visit records between October 2015 and April 2020 (patients aged 12 and older) in which the identified ICD-10 codes had been used in any diagnostic section of the ED visit record. The percentage of “definite” cases seen at each institution ranged from 74.5% (UK) to 26% (UCSD) across the network, suggesting substantial variability in coding practices. Ongoing work is evaluating the accuracy and calibration of the mapped codes and any improvements in categorization afforded by ML methods.

### NLP and ML/AI Development

The NLP data selection and processing steps to generate the development and validation data sets are shown in Figure 4. NER+ Rules identified 1,513 candidate opioid-related OD content sentences from a random 20% sample of 2.47 million clinical notes. These candidate sentences had one or more words or phrases that matched the terms in the NLP opioid-related OD data dictionary, but did not necessarily indicate an OD event or relevant concept. These sentences were manually annotated by reviewers as to either containing (“True” sentences) or not containing (“False” sentences) targeted opioid-related OD content and were then partitioned into three datasets: training (1,213), development (150), and test (150) with equally distributed “True” and “False” cases. In the test dataset, 114 true positive cases and 36 true negative cases (determined via expert chart review) were used to measure the performance of NLP approaches in abstraction of target concepts (Table 2). The results demonstrated that the DL approach, BERT, with post-processing rules achieved the best performance. Future work will examine the portability of these methods across other sites in the consortium.

**Figure 4.**
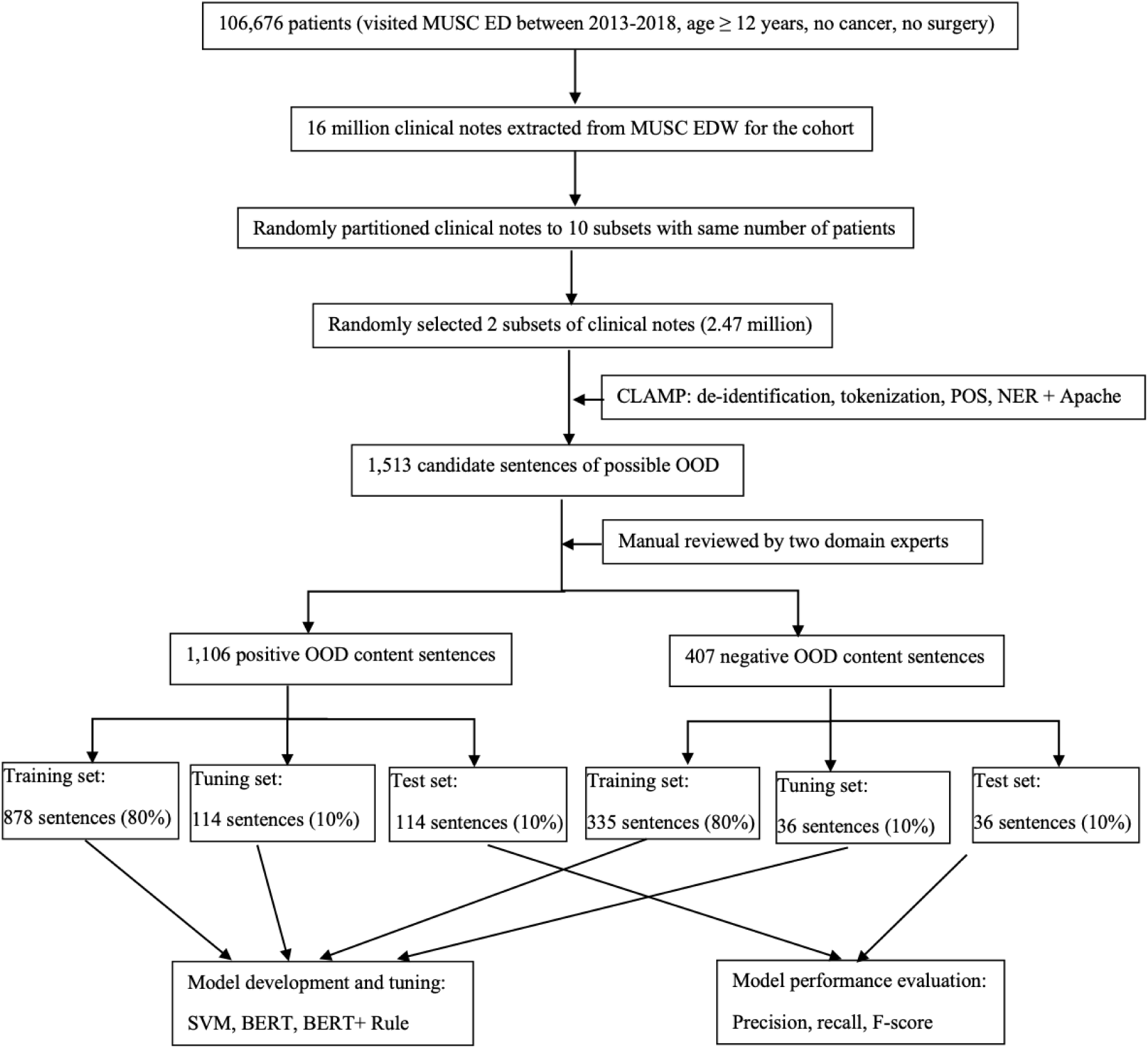
Data processing to develop the test and validation sets for opioid overdose concept abstraction.

**Table 2.** Performance of NLP methods.

| **NLP Approaches** | **Predicted** | **Correct** | **Precision** | **Recall** | **F-score** |
| --- | --- | --- | --- | --- | --- |
| **NER + Rules** | 137 | 105 | 0.77 | 0.92 | 0.84 |
| **SVM** | 122 | 105 | 0.86 | 0.92 | 0.89 |
| **BERT** | 117 | 103 | 0.88 | 0.90 | 0.89 |
| **BERT+Rules** | 106 | 103 | 0.97 | 0.90 | 0.94 |

### EHR documentation tools

The opioid documentation tool shown in Figure 2 was initially available to providers on July 30th, 2020. The OST, plus the “reminder” for use, went into effect on February 25th, 2021. The effect of the reminder to use the OST was assessed by examining the rate of OST use among ED cases with categorized ICD codes, representing a “high probability” of an OOD case (definitions 1 and 2 in Supplemental Table 2) between December 1st, 2020 and April 30th, 2021. Prior to implementation of the reminder, there were zero clinical uses of the OST. There were 43 recorded uses of the OST by 26 different users during the test period. As shown in Table 3, the OST reminder triggered in roughly 21% of cases categorized as high probability (n=81) for OOD during the test period at MUSC Charleston.

**Table 3.** Opioid smart tool “reminder” trigger and use counts (percentages) in high probability cases at MUSC Charleston and within the MUSC regional hospitals.

| **Location** | **MUSC Charleston** | **MUSC Regional Hospitals** |
| --- | --- | --- |
| **High probability cases** | 389 | 292 |
| **Template triggered by real time data** | 81 (21%) | 130 (45%) |
| **Template used for documentation** | 23 (28%) | 18 (14%) |

The MUSC hospital system had recently acquired four regional hospitals, and we were surprised to see that ED providers at the newly acquired regional hospitals were also using the OST, despite not having received any training. In a subset of cases seen in these facilities (n=130), the rate of template use in high probability cases was 14% (Table 3).

An electronic REDCap survey was sent to the 26 providers who used the OST. This survey measured system useability scale (SUS) [18] and net promoter score ratings (NPS) [19]. Twelve providers responded to the survey (46%). The overall SUS score was 72.5% and the NPS score was 74.2% among respondents, suggesting an overall positive evaluation of the OST and the reminder triggering it. Work to implement and test this reminder plus the OST at the other three participating study sites is ongoing.

## DISCUSSION

Investments in research infrastructure need to be adaptable to the evolving demands of society for health research. In this paper we describe our experiences in the first two years of operations, adapting the ACT network to a new use case: surveillance of, and planning for, trials to address the opioid epidemic. The approach has three areas of emphasis that are likely to be important elements of any focused registry like activity: (1) e-phenotype definition and harmonization of coding, (2) enrichment of extent data with NLP of clinical notes, and (3) enhancement of prospective capture of data with informatics-based tools. Each of these areas posed unique challenges.

We developed our platform for the ACT network as it has become the standard for CTSA institutions for planning clinical trials. We recognize that similar enhancements might be made to PCORnet [20] infrastructure, which has a similar sized footprint, or Observational Medical Outcomes Partnership (OMOP) [21] data warehouse infrastructure. There are advantages to taking a canonical data model approach and integrating data production pipelines for various clinical data models upstream [22] but discussion of the canonical approach is beyond the scope of this paper.

E-phenotype definition for OOD posed certain challenges with the degree of its under-reporting in ICD-10 diagnostic and billing codes. Ongoing work is comparing the distribution of codes at each site and will estimate the precision and calibration of categorization using the model across sites.

NLP was found to be an important tool in identifying cases of OOD from text descriptions. Achieving both high precision (.97) and high recall (0.9) for OD cases appears possible using the combination of a BERT model with a rule-based approach. Future work will compare NLP/ML methods with expert derived categories and deep neural network text classifiers, using pre-trained word2vec embedding, to characterize clinical content.

While it is difficult to intervene in busy clinical environments to help clinicians produce “research quality” documentation, the integration of a reminder into the note, triggered by available data at the time of authorship appears to be a potentially valuable tool. The template included links to order sets for care based on guidelines, such as prescribing of naloxone for home use. Thus, it also provides an opportunity to improve the quality of care for OOD as well as documentation. However, triggering insertion of the template requires “real time” data such as the chief complaint, which may not always be present. Additionally, its use requires changing clinician documentation behavior, which is a difficult task. Our hope is that opportunities for improvement in care through implementation of guidelines will help drive adoption of research related documentation.

The work described herein is still at an early stage. While created with dissemination to partners in mind, the actual task of dissemination and assessment of effectiveness remains. Persistent challenges include variability in coding [23,24] and potential differences in the sensitivity and specificity of e-phenotype definitions across institutions sharing a common data model [25]. Access to clinical data for NLP development varies across sites, though all sites were eventually able to enrich project-associated data assets with text notes. Three of four sites used the same EHR system (Epic), while the fourth is converting to Epic. This will facilitate making documentation tools available in each environment and aid in testing the effects of prompting use of the OST. The infrastructure and funding for these next steps is in place.

## Limitations

The question of whether a general purpose network such as ACT can be easily extended, at a low cost, to address targeted and emergent problems remains open. There are many challenges and, as often is true in informatics, the technical challenges are the least significant ones. However, it is clear no single technology in isolation (e-phenotyping, NLP or ED guidelines and templates) can solve all the issues. Only with a combination of approaches is there prospect for success.

## Conclusions

Our initial work suggests it is feasible to extend the ACT network infrastructure, with its real time query capabilities, to begin addressing challenges of response to the opioid pandemic. NLP can play an important role in enhancing derived OUD e-phenotypes. However, developing truly scalable tools that can be deployed across health systems, even those sharing a similar CTSA data warehousing and EHR infrastructure, is a difficult task requiring sustained investment, support for operations and training for clinicians.

## Data Availability

Data in this manuscript are available on written request to the principal investigator.

## Acknowledgements

The authors acknowledge the outstanding project support provided by the data management teams at each of the four CTSAs involved in this project. In addition, we specifically thank Elizabeth Szwast for her outstanding help with management of this complex project.

